# Automated Medical Chart Review for Breast Cancer: A Novel Natural Language Processing Software System

**DOI:** 10.1101/2021.05.04.21256134

**Authors:** Yifu Chen, Lucy Hao, Vito Z Zou, Zsuzsanna Hollander, Raymond T Ng, Kathryn V Isaac

## Abstract

The incoming health records to the BC Cancer Registry are processed between two to three years behind real-time. In response, we developed a Natural Language Processing (NLP) software to automate the electronic chart review workflow. For the same task that costs hundreds of hours of trained labour, our pipeline extracts data within minutes. During preliminary evaluation, an MD student yielded 93.0% and 98.2% accuracies on a sample of operative and pathology breast cancer documents (for a total number of 2,563 data points processed). In comparison, our prototype achieved 89.6% and 91.4% accuracies, respectively. Future plans include improving the performance of the pipeline and eventually adapt it to accepting a more comprehensive range of electronic health records across cancer types and diseases. In the context of BC’s digital healthcare transformation initiatives, this customized software may provide time and cost savings for both the Registry and cancer researchers.

## Introduction

The burden of cancer is recognized as an important challenge in Canadian healthcare due to the growing number of cases annually, morbidity, detrimental impact on quality of life, and cost.^1,2^ Breast cancer is the most common cancer in women, with an estimated 27 000 Canadian women newly diagnosed in 2020.^3^ Breast cancer care is complex and necessitates integrating clinical, pathologic, and imaging data to devise a comprehensive treatment plan. Advances in diagnostics and treatments have improved breast cancer patient outcomes, including survival and quality of life. Translation of clinical research into practice leads to betterment of individuals with cancer. Cancer staging and clinical outcomes data for patients with breast cancer are collected in regional databases and collated in national databases. As the number of patients with cancer increases, a parallel growth occurs in their interactions with the healthcare system and resultant number of medical records. Electronic formats of medical records are becoming ubiquitous across health care systems.^4^ Electronic health records (EHRs) are a fruitful source of information that can be utilized to garner novel understanding of a disease’s natural history,^5^ treatment responses, and prognosis^6–8^ to guide and advance clinical research. In Canada, extraction of this valuable information from EHRs is currently performed by manual review by expert data extractors and clinical researchers, which significantly limits timeliness, scalability, and research due to high costs.

Regional Cancer Registries (databases) are mandated to report on cancer outcomes and support oncologic population health research. To assure a high level of data quality and completeness, input of clinical data on cancer staging and outcomes is by manual extraction from patient charts; this method of data extraction is time consuming, costly, and inefficient. Currently, the volume of work for data extraction exceeds capacity, leading to time delays and restrictions on the scope of variables extracted. The BC Cancer Registry is processing incoming data at two to three years behind real time, and this gap is widening at approximately 3% per year. This is partially due to the increased volume and complexity of data rising with the synchronous expansion of cancer cases and clinical knowledge pertinent for diagnosis, management, and prognosis. With finite resources in the Canadian health care system, regional cancer databases must limit and prioritize the extraction of specific variables of known relevance to treatment planning and prognosis. In response to strained resources, the scope of information and timeliness of data input into cancer databases have suffered and are limited as opposed to expanding.

Computational methods to automate and expedite data extraction from medical records are rapidly developing as an expanding field in computer science. Natural Language Processing utilizes computational methods to analyzed language, where text and speech data inputs are used for processing and capturing meaning from words. NLP algorithms are most commonly tasked to extract text and recognize specific entities. Current methods for text processing in the cancer domain include three relevant categories of NLP strategies: named entity recognition (NER), information extraction (IE), and text classification. NER identifies terms and classifies these according to predefined categories, relying on dictionaries of biomedical terms, for e.g. Unified Medical Language System (UMLS) metathesaurus used via MetaMap to obtain terms annotated to entities.^9^ An important challenge with reliance on standardized dictionaries is the common existence of term variability. Terms may often not be found in the source dictionaries because of synonyms, acronyms, abbreviations, or idiosyncrasies (like grammatical errors) which requires additional strategies.^10^ IE methods identify predefined facts and relationships of interest, often using NER and additional modelling using regular expression pattern-matching rules and negation rules. This fine-tuning approach is highly reliable for IE of valuable clinical information specific to a cancer type (ie. Nottingham score for breast cancer), especially when records have structural conformity.^11^ Text classification extends the benefits of IE to infer information that is not explicitly stated, but derived by predefined rules, whereby a cancer can be classified into a predefined category according to an expert-derived program of rules for inductive reasoning.^12^ With the hierarchical building of IE and TC strategies on foundational methods of NER, it is critical to have high accuracy in the lexicon. Any baseline errors or compromise in the entities upon which the final algorithm is built will limit the scalability, accuracy, and generalizability of the algorithm.

To accomplish both the national goals of obtaining prespecified cancer staging and clinical research goals of obtaining prespecified surgical and cancer outcomes variables, the research team explored existing NLP software tools that were widely used to process similar data. Ashish et al developed enhancements to the academic institutional information extraction system to expand the data fields of interest for automated capture of data from cancer pathology reports.^13^ This pipeline was built leveraging the existing Unstructured Information Management Architecture (UIMA) framework and resources from the Open Health Natural Language Processing (OHNLP) consortium. Their Pathology Extraction Pipeline is built upon the established Medical Knowledge Analysis Tool pipeline (MedKATp), which focuses on pathology reports. Xie et al utilized the Text Information Extraction System (TIES) to identify potential new cancer diagnoses in real-time using concept terms from the National Cancer Institute Metathesaurus (NCIM) and codes to identify breast cancer from the Unified Medical Language System Terminology Service (UTS). However, these software solutions were deemed inadequate due to rigid algorithms that lack the capacity to customize formatting and scripts. With the rapidity of novel computational method development, it is essential to ensure the programming of the algorithm can be updated with advances both in NLP strategies and in clinical research. Adapting and extending the existing tools is very technically demanding and still cannot guarantee that it is both customizable to meet BC standards and extensible to ensure long-term usability.

To address these challenges, we developed a fully customized NLP algorithm as a solution to our goal of automating extraction of clinically relevant diagnostic, treatment, and prognostic variables into a population-based cancer database for a regional jurisdiction. This de novo rule-based, transparent, and explainable algorithm runs a uniquely generated “pattern matcher” of custom rules to extract a given set of predefined clinical variables of interest. We then encode the extractions with a biomedical text NLP model that was trained on large-scale biomedical datasets.^14^ The algorithm development was based on a curated sample of 100 breast pathology and operative notes each. These notes contain a standardized “Synoptic Report” section, and we leveraged its structural conformity to develop the customized NLP algorithm.

## Methods

### Task Description

The retrospective chart review (RCR) is a task that extracts data variables from pre-recorded health data. Human reviewers carefully read and examine each clinical note to extract and encode target columns of interest onto a spreadsheet. A codebook standardizes the extraction of numeric encodings. In our study, the operative and pathology reports contain 8 and 37 extractable columns, respectively. Medical students manually extracted 100 operative and 100 pathology reports to establish the baseline performance. It was apparent that the chart review process is very labor intensive – a trained MD student spends about 20 minutes on each clinical note document, and for large-scale retrospective studies, thousands of such medical records may be used.

### NLP Pipeline Overview

The NLP pipeline is engineered with the end-to-end capacity to substitute manual reviewers. It takes a list of PDF health records and outputs the numeric encodings on an Excel spreadsheet. A high-level diagram of this pipeline is shown in Figure 1. In overview: it first reads the entire document using Optical Character Recognition (OCR) and searches for a “Synoptic Report” section. Then it identifies the key variables and encodes the information. In cases the variable is not easily identifiable, the software expands the search criteria while autocorrecting any potential typos in column names. The software outputs the encoded and uncoded variable extractions in the final stage.

**Figure 1.**
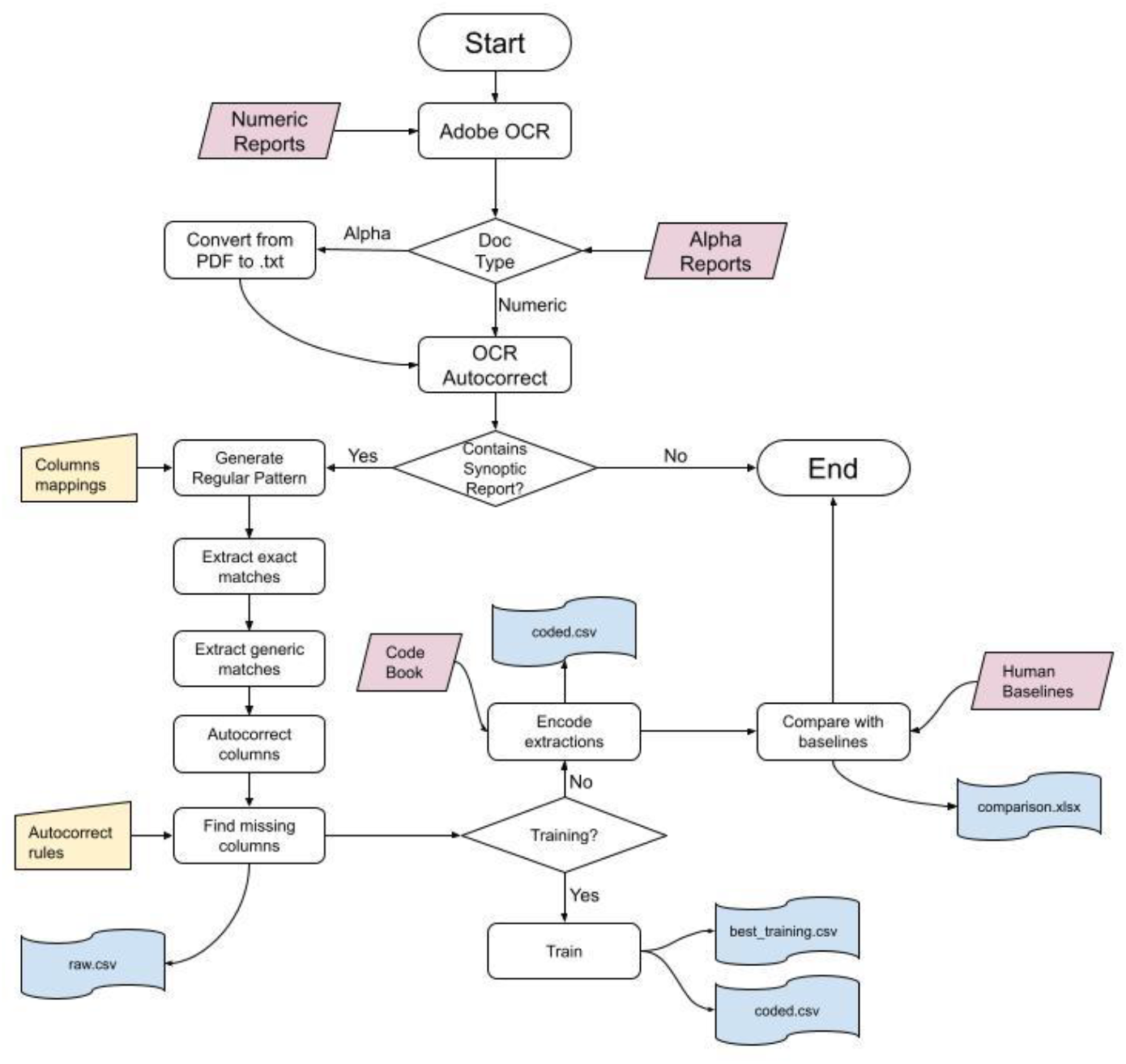
Flowchart of pipeline process

**Figure 2:**
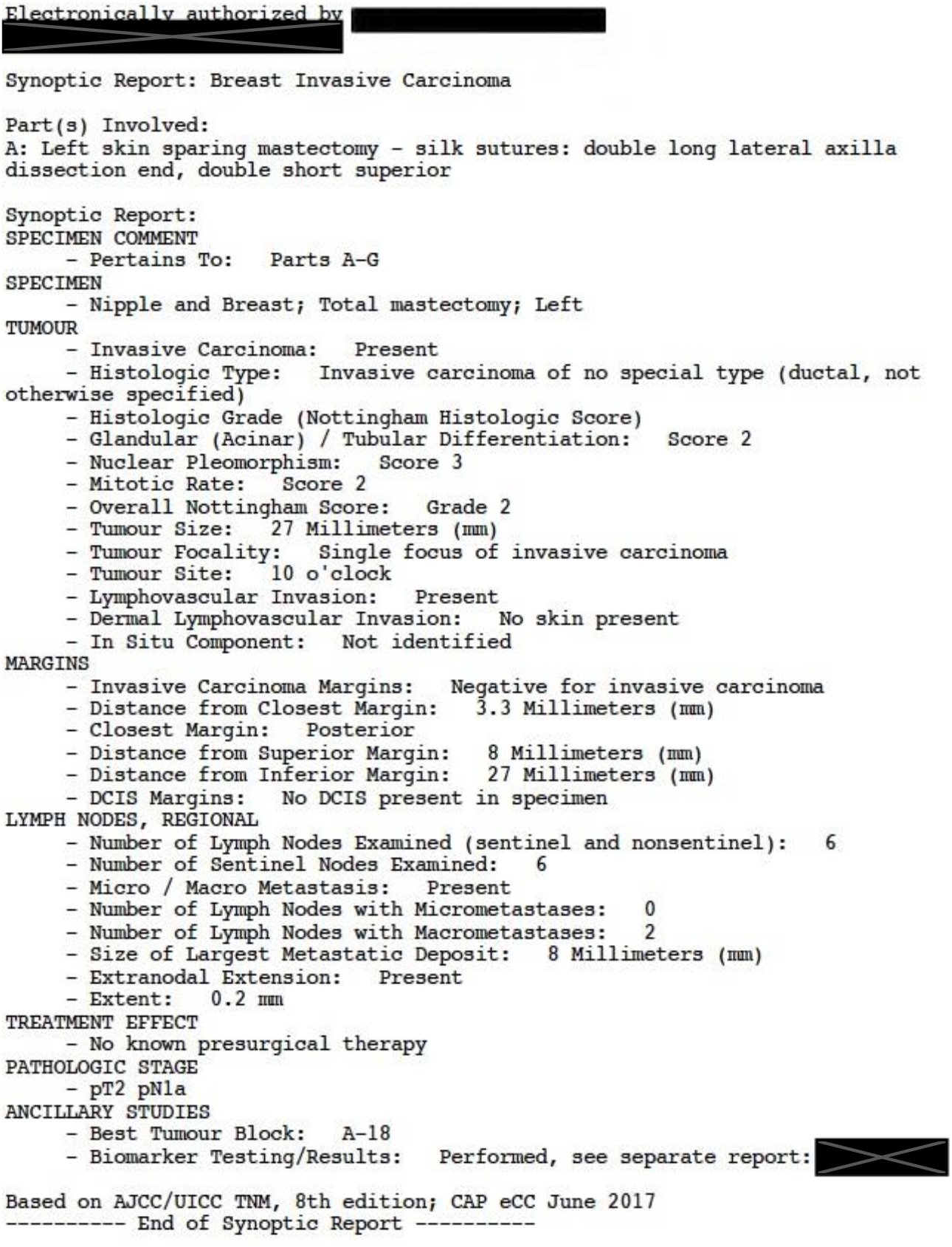
A Synoptic Report contains structured point-form data that summarizes the report.

We combined custom algorithms with peer-reviewed biomedical text processing and computer vision research.^14,15^ The pipeline has three main advantages compared to alternative software products: 1) full customizability of the codebook encodings, and 2) easy extensibility for future development, which allows 3) programmers learn from “past mistakes” and quickly improve the extraction accuracy through reprocessing all documents.

### Implementation Details

#### Preprocessing

The pipeline consists of several pre-processing modules to prevent errors that occur when the input documents are in unideal formats. There are two different methods to preprocess the PDFs, depending on their content. For reports that mainly contain alphabetical values (operative reports), pre-processing of the PDF is included as a module within the pipeline. An open-source OCR tool, pytesseract,^15^ is used to transform PDFs to images in which text can be recognized and read. For pathology reports, Optical Character Recognition (OCR) is performed using Adobe Acrobat’s OCR to extract the text data of the scanned PDF.

Pytesseract offers significant accuracy gain compared to Adobe Acrobat on reports with mainly alphabetical values; it is very accurate, leaving almost no misinterpreted characters. However, pytesseract does not handle well texts containing numerical values. Therefore, for pathology reports with many numerical values to extract (e.g. tumour size, distance from margins, etc.), we use the out-of-box tool, Adobe Acrobat. However, one common error due to Adobe Acrobat is word fragmentation. Another module will improve the text recognition by resolving any extra or missing white spaces from OCR. We also run auto-correction for each bullet point.

The Graphical User Interface of our pipeline will display a table for the auto-corrected text for clinician review. If the auto-correction was not supposed to be performed, the clinician is able to flag these errors and re-run the system to produce the corrected result within minutes.

#### Processing

After preprocessing is complete, the pipeline begins extracting the encodings for the Fields of Interest (FoI) listed in the codebook. To make the pipeline extendable to other types of synoptic reports, there is a regular pattern matcher generation feature. Based on the FoIs, a regular pattern is generated. This regular pattern can be fine-tuned in a variety of ways.

In case any of the FoIs are not captured by the generated regular pattern (this can happen if the column has a slight misspelling or there are white spaces), there is a second regular pattern which looks for the separator (usually a colon “:”) and captures all column value pairs with that separator.

Since some column values can help determine other column’s value, we also applied some rules to the pipeline through custom functions. For instance, if “No lymph nodes present” is in the report, then the pipeline specifies values for the two FoIs, “number of lymph nodes with micro-metastases” and “number of lymph nodes with macro-metastases”, as None.

#### Postprocessing

After the values have been extracted, we extract entity encodings from values using scispaCy, which are spaCy models trained on biomedical text (Neumann 2019). If the value is shorter than 3 words, we did not extract entities. This is because for some terms like “wise pattern”, there are no extracted entities. We use the “en_core_sci_lg” model which has a vocabulary size of 785,000. After the entities are extracted, we tokenize using spacy and use cosine similarity to choose the closest encoding from our code book. For an encoding to be returned, a similarity threshold must be passed. If the similarity cannot pass the threshold, the raw extracted value is returned instead of an encoding. For each FoI, this threshold is determined in the training period of the pipeline.

In the training period of the pipeline, we try a range of thresholds and find the best threshold for each FoI. We keep the lowest threshold and the highest threshold that produce the same results. For instance, if a threshold of 0.85 produces the same accuracy for a FoI as 0.95, we average the thresholds to get 0.90. See Appendix E for an example of training thresholds.

#### Extraction Visualization

Another unique custom feature is the visualization tool that displays the comparison of results between the Excel sheets produced by a clinician versus the pipeline. The entries are highlighted in yellow, red, and green according to whether the pipeline extracted a value that is different, missing, or extra compared to the human-extracted spreadsheet. An example is displayed in Figure 3. This feature enables researchers and clinicians to cross-check the manual extraction with the software results.

**Figure 3:**
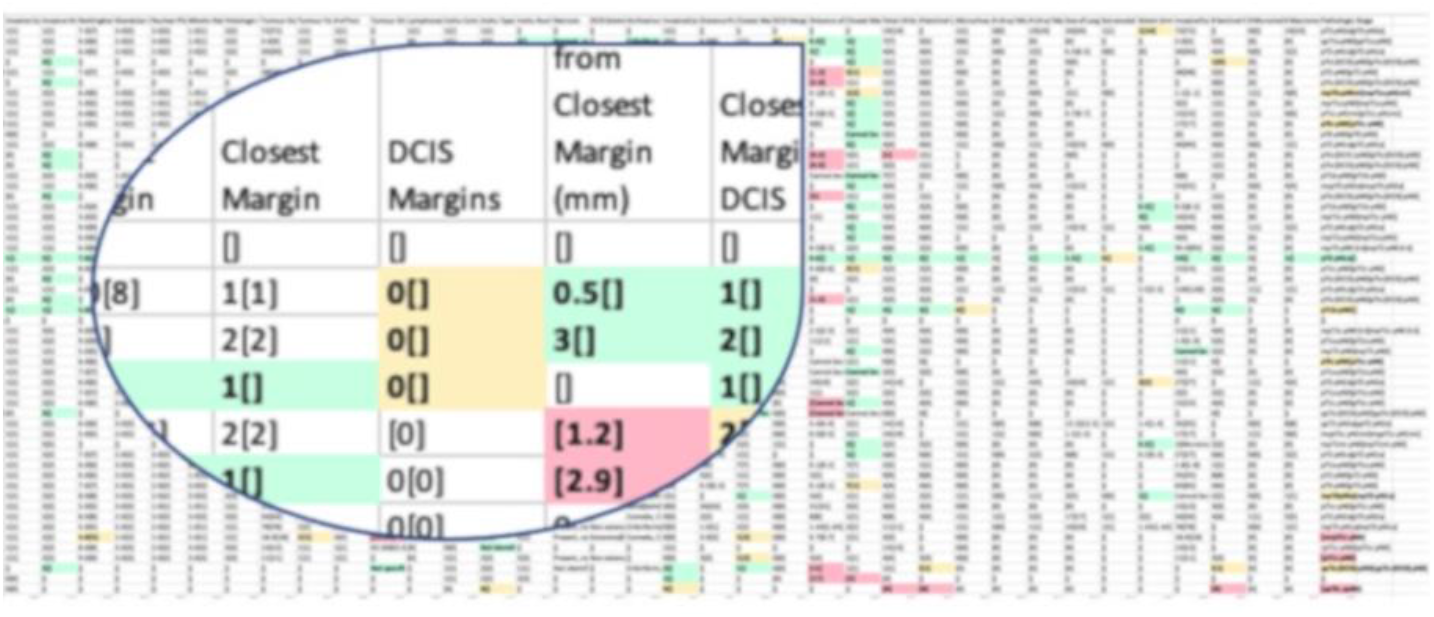
A zoomed-in view of the comparison between human and machine extractions.

Therefore, in addition to the possible scenario of substituting the manual reviewers completely, this pipeline may initially serve as a result checker for clinicians. This has a win-win effect – while manual chart reviewers address potential mistakes or gain confidence in the double-checked extractions, the software team also learns to fine-tunes the software to prevent any errors from occurring again in the future. Given enough data and time, this iterative development model allows the pipeline accuracy to asymptotically approach human accuracies.

## Results

The overall accuracy across the 200 total reports is found to be 90.4%. The operative and pathology report accuracies are 89.6% and 91.4%. In comparison, a medical student achieved accuracies of 93.0% and 98.2% respectively. We calculated the additional detailed metrics for the each FoI, as shown in the Appendices C, D, E, F, and G.

Given the high levels of accuracy, we believe that our pipeline is a promising validator (and potentially a substitution) to the manual extraction by clinicians. While our medical student spent over a hundred hours to finish processing the 50 pathology reports and 50 operative reports, it takes just a few minutes when our pipeline is operated by a human (although some additional time could be spent to review and correct the auto-corrections). The estimated computation time required to classify tens of thousands of documents is a few hours if the pipeline is run on a normal laptop, in comparison to many months of person-hour. Since the pipeline outputs an additional spreadsheet for all variables other than FoIs too, we could rapidly extract information when new FoIs are added in the future. We plan to conduct a thorough error analysis and review by clinicians, but some additional observations at this time are detailed below.

One column that was surprisingly low was the Insitu Type column. Our training accuracy was 96% while our validation accuracy was 75%. After further inspection, we realize this error was due to the change in validation baselines and our treatment of extra values. Sometimes the pipeline will extract extra information that is not covered in the code book. We chose to keep those extra values seeing that they could provide some value and did not penalize the pipeline for extra values. When there is no Insitu Type, the value “Invasive carcinoma of no special type (ductal, not otherwise specified),” is extracted. Our pipeline extracts an entity of “ductal” and wrongly classifies it as DCIS. Our training baselines did not input any encoding for this value and left it blank, while our validation baselines encoded a value of 0 for “Invasive carcinoma of no special type”. Thus, not specifying an encoding in the training set caused the pipeline to wrongfully return information. In the future, for a robust pipeline, the human encoders must be consistent in how they deal with negative values.

Thus, some columns may never be “100%” accurate due to the subjective interpretation of each surgeon. We saw this occur for Incision Type many times. When an incision removes the entire tumor, a mastectomy is done. When a mastectomy is done and the entire tumor is removed, surgeons do not need to keep track of what incision was used because they will likely not need to reopen that incision. However, if only a partial mastectomy was completed, the surgeon may need to go back and open the incision, as the entire tumor was not extracted. Even in the case of a complete mastectomy, some surgeons may still want to know what incision type was used. This was seen in how our human encoders extracted the incision types; some extracted the incision type no matter what type of mastectomy was performed, while another extractor only encoded the incision type if it was directly stated right beside “Incision type and its relation to tumor”.

## Discussion

Some disadvantages, however, include the following. If any report slightly deviates from this synoptic style, our regular pattern may not extract the entire report. Misspellings or spurious white spaces can cause the regular pattern not to recognize the column. In response, we created a mechanism, which auto-corrects the column names and displays the autocorrection to clinicians for review (an example is available in Appendix F). Finally, if some columns are reported with a slight variation, (e.g., “incision and incision relation to tumor” vs. “incision and its relation to tumor”) our regular pattern cannot determine that these two columns are in fact the same column, but if the columns are close enough (depending on our autocorrect algorithm) they are still matched as similar. Lastly, with a general biomedical text NLP model and small training set, our encoding algorithm sometimes makes mistakes – but this can be mitigated with a larger training set or more customized NLP model.

Another limitation of the pipeline is that is that it cannot intelligently understand the report. Many of the columns in the pathology report are not completely independent, meaning the value in one column can help determine the values in other columns. For instance, if the report mentions “Negative for DCIS”, some FoIs should be left blank. For now, the user must explicitly specify this into the pipeline; the pipeline cannot learn this behavior. The advantage is that if a user is certain of a behavior, they can quickly make the pipeline adopt this behaviour through a custom function; there is no need to train for hours. We were able to determine some of these relationships between FoI in the training set if accuracies of certain FoI in the training set were low to start with. For instance, the accuracies for Closest Margin, DCIS Margins, Distance of DCIS from Closest Margin (mm) and Closest Margin DCIS were very low without the custom rules. After a custom rule was added for these FoIs, the accuracy increased by 10+% in the training set. In the validation set, two columns were able to retain their training accuracy.

A portion of our sample of reports do not contain the “Synoptic Report” section. Without this section, the current pipeline cannot perform the data extraction task with sufficient levels of accuracy. In response, we are currently developing additional methods by using “Final Diagnosis” section as the data source. The Final Diagnosis section contains semi-structured text which span multiple paragraphs. We plan to use a machine learning Natural Language Processing parser to construct a dependency relations tree. With the dependency relations, we can extract location-specific labels for the FoIs.

## Conclusion

We decided to create a fully custom pipeline with custom algorithms to be able to easily add and remove FoIs. For instance, if any researcher decides to extract 5 more columns, instead of necessitating the re-extraction for all documents, they simply need to specify add columns to a pipeline configuration Excel file. We took advantage of the unified structured in the synoptic sections by utilizing regular patterns matching. We use a custom algorithm to flexibly identify and generate a unique regular pattern based on the Synoptic Report template. Thus, our regular pattern generation algorithm can be tuned within minutes and reprocess thousands of documents easily. Since we did not have access to thousands of reports, this also limits the types of NLP methods based on machine learning; the advantage with regular patterns is that we can create a rule from just a few reports if we know most reports will follow the same format. Similarly, we decided to create our own encoding algorithm with use of general biomedical entity extraction model to have this same flexibility in adding new encodings to our code book. As result, our pipeline is fully rule-based, transparent, and interpretable which is critical in clinical applications.

## Data Availability

Data is available upon request from the authors.

## Acknowledgements

The authors would like to thank Shawn Yuan and Dorsa Mousa-Doust for their assistance with medical chart review.

