## Appendices for "Automated Medical Chart Review for Breast Cancer: A Novel Natural Language Processing Software System"

```
Synoptic Report: Breast Invasive Carcinoma

Part(s) Involved:
A: right breast - long lateral,short superior

Synoptic Report:
SPECIMEN COMMENT
     - Pertains To:   Specimen A to C
SPECIMEN
     - Breast; Total mastectomy; Right
TUMOUR
     - Invasive Carcinoma:   Present
     - Histologic Type:   Invasive carcinoma of no special type (ductal,
not otherwise specified)
     - Histologic Grade (Nottingham Histologic Score)
     - Glandular (Acinar) / Tubular Differentiation:   Score 3
     - Nuclear Pleomorphism:   Score 3
     - Mitotic Rate:   Score 3
     - Overall Nottingham Score:   Grade 3
     - Tumour Size:   1.9 Millimeters (mm)
     - Tumour Focality:   Single focus of invasive carcinoma
     - Tumour Site:   10 o'clock
     - Lymphovascular Invasion:   Not identified
     - In Situ Component:   Present
     - In Situ Component Type:   DCIS
     - Nuclear Grade:   Grade III (high)
     - Necrosis:   Present, focal (small foci or single cell necrosis)
     - DCIS Extent:   Non extensive
     - DCIS Estimated Size:   19 mm
     - Architectural Patterns:   Cribriform, Solid
MARGINS
     - Invasive Carcinoma Margins:   Negative for invasive carcinoma
     - Distance from Closest Margin:   11 Millimeters (mm)
     - Closest Margin:   Posterior
     - DCIS Margins:   Negative for DCIS
     - Distance of DCIS from Closest Margin:   11 Millimeters (mm)
     - Closest Margin:   Posterior
LYMPH NODES, REGIONAL
     - Number of Lymph Nodes Examined (sentinel and nonsentinel):   5
     - Number of Sentinel Nodes Examined:   5
     - Micro / Macro Metastasis:   Not identified
     - Number of Lymph Nodes with Isolated Tumour Cells:   0
PATHOLOGIC STAGE
     - pT1a pN0
ANCILLARY STUDIES
     - Best Tumour Block:   A4
     - Biomarker Testing/Results:   Performed, see separate report:
[redacted]

Based on AJCC/UICC TNM, 8th edition; CAP eCC June 2017
---------- End of Synoptic Report ----------
```

Appendix A: Sample Synoptic Pathology Report

| Metric | (=) | Formula |
|---|---|---|
| Precision | PPV | $\frac{TP}{TP+FP}$ |
| Recall | Sensitivity | $\frac{TP}{TP+FN}$ |
| F-Score | F | $\frac{TP}{TP+\frac{1}{2}(FP+FN)}$ |
| Specificity | | $\frac{TN}{TN+FP}$ |
| Net Present Value | NPV | $\frac{TN}{TN+FN}$ |
| Accuracy | | $\frac{TP+TN}{TP+TN+FP+FN}$ |

Appendix B: Metric formulae used in appendices C, D, E, and F

| | Invasive Carcinoma | Invasive Histologic Type | Nottingham Score | Glandular Differentiation | Nuclear Pleomorphism | Mitotic Rate | Histologic Grade | Tumour Size (mm) | Tumour Focality | # of Foci | Tumour Site | Lymphovascular Invasion | Insitu Component |
|---|---|---|---|---|---|---|---|---|---|---|---|---|---|
| Same | 50 | 42 | 49 | 49 | 50 | 49 | 50 | 49 | 48 | 51 | 39 | 51 | 50 |
| Different | 2 | 0 | 2 | 1 | 0 | 1 | 0 | 0 | 2 | 0 | 1 | 1 | 1 |
| Missing | 1 | 1 | 1 | 1 | 1 | 1 | 1 | 3 | 1 | 1 | 0 | 0 | 1 |
| Extra | 0 | 10 | 1 | 2 | 2 | 2 | 2 | 1 | 2 | 1 | 13 | 1 | 1 |
| Accuracy | 0.94 | 0.98 | 0.94 | 0.96 | 0.98 | 0.96 | 0.98 | 0.94 | 0.94 | 0.98 | 0.97 | 0.98 | 0.96 |

| | Insitu Type | Insitu Nuclear Grade | Necrosis | DCIS Extent | Archtectural Patterns | InvasiveCarcinoma Margins | Distance from Closest Margin | Closest Margin | DCIS Margins | Distance of DCIS from Closest Margin (mm) | Closest Margin DCIS | Total LN Examined |
|---|---|---|---|---|---|---|---|---|---|---|---|---|
| Same | 43 | 50 | 48 | 51 | 50 | 50 | 48 | 45 | 39 | 44 | 45 | 51 |
| Different | 1 | 0 | 1 | 1 | 1 | 3 | 0 | 1 | 5 | 1 | 1 | 1 |
| Missing | 1 | 1 | 1 | 1 | 0 | 0 | 4 | 1 | 0 | 7 | 6 | 1 |
| Extra | 8 | 2 | 3 | 0 | 2 | 0 | 1 | 6 | 9 | 1 | 1 | 0 |
| Accuracy | 0.96 | 0.98 | 0.96 | 0.96 | 0.98 | 0.94 | 0.92 | 0.96 | 0.89 | 0.85 | 0.87 | 0.96 |

| | # Sentinel LN Examined | Micro/macro metastasis | # LN w/ Micrometastasis | # LN w/ Macrometastasis | Size of Largest Macrometastasis Deposit | Extranodal Extension | Extent (mm) | InvasiveTumourSize (mm) | # Sentinel Nodes Examined | # Micrometastatic Nodes | # Macrometastatic Nodes | Pathologic Stage |
|---|---|---|---|---|---|---|---|---|---|---|---|---|
| Same | 51 | 52 | 53 | 52 | 52 | 52 | 47 | 50 | 50 | 53 | 52 | 50 |
| Different | 1 | 1 | 0 | 0 | 0 | 1 | 0 | 0 | 1 | 0 | 0 | 0 |
| Missing | 1 | 0 | 0 | 0 | 0 | 0 | 0 | 2 | 1 | 0 | 0 | 2 |
| Extra | 0 | 0 | 0 | 1 | 1 | 0 | 6 | 1 | 1 | 0 | 1 | 1 |
| Accuracy | 0.96 | 0.98 | 1 | 1 | 1 | 0.98 | 1 | 0.96 | 0.96 | 1 | 1 | 0.96 |

| | Invasive Carcinoma | Invasive Histologic Type | Nottingham Score | Glandular Differentiation | Nuclear Pleomorphism | Mitotic Rate | Histologic Grade | Tumour Size (mm) | Tumour Focality | # of Foci | Tumour Site | Lymphovascular Invasion |
|---|---|---|---|---|---|---|---|---|---|---|---|---|
| same | 49 | 41 | 49 | 50 | 51 | 51 | 50 | 48 | 51 | 49 | 51 | 52 |
| different | 3 | 12 | 2 | 1 | 1 | 1 | 1 | 2 | 1 | 0 | 2 | 1 |
| missing | 1 | 0 | 2 | 2 | 1 | 1 | 2 | 3 | 1 | 4 | 0 | 0 |
| extra | 0 | 0 | 0 | 0 | 0 | 0 | 2 | 1 | 3 | 2 | 6 | 1 |
| Accuracy | 0.92 | 0.77 | 0.92 | 0.94 | 0.96 | 0.96 | 0.94 | 0.91 | 0.96 | 0.92 | 0.96 | 0.98 |

| | Insitu Component | Insitu Type | Insitu Nuclear Grade | Necrosis | DCIS Extent | Archtectural Patterns | InvasiveCarcinoma Margins | Distance from Closest Margin | Closest Margin | DCIS Margins | Distance of DCIS from Closest Margin (mm) | Closest Margin DCIS |
|---|---|---|---|---|---|---|---|---|---|---|---|---|
| same | 50 | 40 | 50 | 50 | 33 | 52 | 51 | 44 | 47 | 40 | 45 | 44 |
| different | 3 | 12 | 2 | 2 | 3 | 1 | 2 | 1 | 1 | 13 | 2 | 2 |
| missing | 0 | 1 | 1 | 1 | 17 | 0 | 0 | 8 | 5 | 0 | 6 | 7 |
| extra | 1 | 3 | 1 | 0 | 0 | 0 | 2 | 1 | 4 | 5 | 2 | 3 |
| Accuracy | 0.94 | 0.75 | 0.94 | 0.94 | 0.62 | 0.98 | 0.96 | 0.83 | 0.89 | 0.75 | 0.85 | 0.83 |

| | Total LN Examined | # Sentinel LN Examined | Micro/macro metastasis | # LN w/ Micrometastasis | # LN w/ Macrometastasis | Size of Largest Macrometastasis Deposit | Extranodal Extension | Extent (mm) | InvasiveTumourSize (mm) | # Sentinel Nodes Examined | # Micrometastatic Nodes | # Macrometastatic Nodes | Pathologic Stage |
|---|---|---|---|---|---|---|---|---|---|---|---|---|---|
| same | 50 | 51 | 46 | 53 | 52 | 52 | 51 | 53 | 49 | 49 | 53 | 52 | 49 |
| different | 2 | 2 | 7 | 0 | 1 | 0 | 2 | 0 | 2 | 4 | 0 | 1 | 3 |
| missing | 1 | 0 | 0 | 0 | 0 | 1 | 0 | 0 | 2 | 0 | 0 | 0 | 1 |
| extra | 1 | 1 | 0 | 0 | 0 | 2 | 0 | 5 | 4 | 4 | 1 | 0 | 0 |
| Accuracy | 0.94 | 0.96 | 0.87 | 1 | 0.98 | 0.98 | 0.96 | 1 | 0.92 | 0.92 | 1 | 0.98 | 0.92 |

Appendix C: FoI-specific Training (top) and Validation (bottom) Accuracies for Pathology Reports

| | Laterality | Surgical Indication | Pre-Operative Biopsy | Pre-Operative Diagnosis | Neoadjuvant Treatment |
|---|---|---|---|---|---|
| Same | 48 | 50 | 50 | 49 | 50 |
| Different | 2 | 0 | 0 | 1 | 0 |
| Missing | 0 | 0 | 0 | 0 | 0 |
| Extra | 0 | 0 | 0 | 0 | 0 |
| Accuracy | 0.96 | 1 | 1 | 0.98 | 1 |
| Breast Procedure | Immediate Reconstruction Mentioned | Immediate Reconstruction Type | Wire Localization | Breast Incision Type | Axillary Surgery |
| 48 | 48 | 40 | 46 | 32 | 49 |
| 2 | 2 | 8 | 4 | 10 | 1 |
| 0 | 0 | 1 | 0 | 5 | 0 |
| 0 | 0 | 1 | 0 | 3 | 0 |
| 0.96 | 0.96 | 0.82 | 0.92 | 0.68 | 0.98 |

| | Laterality | Surgical Indication | Pre-Operative Biopsy | Pre-Operative Diagnosis | Neoadjuvant Treatment |
|---|---|---|---|---|---|
| Same | 46 | 47 | 49 | 47 | 49 |
| Different | 3 | 2 | 0 | 2 | 1 |
| Missing | 1 | 1 | 1 | 1 | 0 |
| Extra | 0 | 0 | 0 | 0 | 0 |
| Accuracy | 0.92 | 0.94 | 0.98 | 0.94 | 0.98 |
| Breast Procedure | Reconstruction Mentioned | Reconstruction Type | Wire Localization | Breast Incision Type | Axillary Surgery |
| 40 | 46 | 39 | 44 | 39 | 47 |
| 10 | 4 | 10 | 5 | 9 | 1 |
| 0 | 0 | 1 | 1 | 2 | 2 |
| 0 | 0 | 0 | 0 | 12 | 1 |
| 0.8 | 0.92 | 0.78 | 0.88 | 0.78 | 0.94 |

Appendix D: FoI-specific Training (top) and Validation (bottom) Accuracies for Operative Reports

| | Invasive Carcinoma | Invasive Histologic Type | Nottingham Score | Glandular Differentiation | Nuclear Pleomorphism | Mitotic Rate | Histologic Grade | Tumour Size (mm) | Tumour Focality | # of Foci | Tumour Site | Lymphovascular Invasion |
|---|---|---|---|---|---|---|---|---|---|---|---|---|
| Precision | 0.953488372 | 0.76 | 0.972972973 | 1 | 1 | 1 | 1 | 0.945945946 | 0.926829268 | 0.970588235 | 0.666666667 | 0.95 |
| Recall | 1 | 1 | 0.972972973 | 0.975609756 | 1 | 1 | 0.972222222 | 0.945945946 | 1 | 0.916666667 | 1 | 1 |
| F | 0.976190476 | 0.863636364 | 0.972972973 | 0.987654321 | 1 | 1 | 0.985915493 | 0.945945946 | 0.962025316 | 0.942857143 | 0.8 | 0.974358974 |

| | Insitu Component | Insitu Type | Insitu Nuclear Grade | Necrosis | DCIS Extent | Archtectural Patterns | InvasiveCarcinoma Margins | Distance from Closest Margin | Closest Margin | DCIS Margins | Distance of DCIS from Closest Margin (mm) | Closest Margin DCIS |
|---|---|---|---|---|---|---|---|---|---|---|---|---|
| Precision | 0.895833333 | 0.68 | 0.902439024 | 0.918918919 | 1 | 0.888888889 | 0.9 | 0.909090909 | 0.888888889 | 0.693877551 | 0.88 | 0.904761905 |
| Recall | 1 | 0.971428571 | 1 | 1 | 0.36 | 1 | 1 | 0.810810811 | 0.888888889 | 1 | 0.814814815 | 0.76 |
| F | 0.945054945 | 0.8 | 0.948717949 | 0.957746479 | 0.529411765 | 0.941176471 | 0.947368421 | 0.857142857 | 0.888888889 | 0.819277108 | 0.846153846 | 0.826086957 |

| Total LN Examined | # Sentinel LN Examined | Micro/macro metastasis | # LN w/ Micrometastasis | # LN w/ Macrometastasis | Largest Macrometastasis Deposit | Extranodal Extension | Extent (mm) | InvasiveTumourSize (mm) | # Sentinel Nodes Examined | # Micrometastatic Nodes | # Macrometastatic Nodes | Pathologic Stage |
|---|---|---|---|---|---|---|---|---|---|---|---|---|
| 0.938776 | 0.933333333 | 0.85106383 | 1 | 0.954545455 | 0.833333333 | 0.846153846 | 0.428571429 | 0.846153846 | 0.808510638 | 0.909090909 | 0.952380952 | 0.959183673 |
| 0.978723 | 1 | 1 | 1 | 1 | 0.909090909 | 1 | 1 | 0.970588235 | 1 | 1 | 1 | 0.979166667 |
| 0.958333 | 0.965517241 | 0.91954023 | 1 | 0.976744186 | 0.869565217 | 0.916666667 | 0.6 | 0.904109589 | 0.894117647 | 0.952380952 | 0.975609756 | 0.969072165 |

Appendix E: Statistics for Pathology Reports

| | Laterality | Surgical Indication | Pre-Operative Biopsy | Pre-Operative Diagnosis | Neoadjuvant Treatment |
|---|---|---|---|---|---|
| Precision | 0.93877551 | 0.979166667 | 1 | 0.96 | 0.980392157 |
| Recall | 0.978723404 | 0.979166667 | 0.98 | 1 | 1 |
| F | 0.958333333 | 0.979166667 | 0.98989899 | 0.979591837 | 0.99009901 |

| Breast Procedure | Immediate Reconstruction Mentioned | Immediate Reconstruction Type | Wire Localization | Breast Incision Type | Axillary Surgery |
|---|---|---|---|---|---|
| 0.854166667 | 0.921568627 | 0.795918367 | 0.897959184 | 0.45 | 0.979166667 |
| 1 | 1 | 0.975 | 0.977777778 | 0.9 | 0.959183673 |
| 0.921348315 | 0.959183673 | 0.876404494 | 0.936170213 | 0.6 | 0.969072165 |

Appendix F: Statistics for Operative Reports

| | A | B | C | D | E | F | G | H |
|---|---|---|---|---|---|---|---|---|
| 1 | | column | t best thre | t best thre | threshold | same | extra | |
| 2 | **0** | Laterality | 0.7 | 0.95 | 0.82 | 48 | 0 | |
| 3 | **1** | Surgical Ir | 0.7 | 0.8 | 0.75 | 50 | 0 | |
| 4 | **2** | Pre-Opera | 0.7 | 0.9 | 0.8 | 50 | 0 | |
| 5 | **3** | Pre-Opera | 0.7 | 0.95 | 0.82 | 49 | 0 | |
| 6 | **4** | Neoadjuv | 0.7 | 0.95 | 0.82 | 50 | 0 | |
| 7 | **5** | Breast Prc | 0.7 | 0.9 | 0.8 | 48 | 0 | |
| 8 | **6** | Immediat | 0.7 | 0.95 | 0.82 | 39 | 1 | |
| 9 | **7** | Wire Loca | 0.7 | 0.95 | 0.82 | 46 | 0 | |
| 10 | **8** | Breast Inc | 0.8 | 0.8 | 0.8 | 32 | 3 | |
| 11 | **9** | Axillary Su | 0.7 | 0.9 | 0.8 | 49 | 0 | |
| 12 | **10** | Immediat | 0.7 | 0.7 | 0.7 | 47 | 0 | |
| 13 | | | | | | | | |

Appendix G: Example of thresholds when using vector similarities for encoding text into numbers

Double-click any column header to sort the table

| | Study ID | Original Column | Corrected Column | Edit Distan | Extracted Data |
|---|---|---|---|---|---|
| 1 | 18 | incision in relation to the tumor | incision in relation to tumor | 4 | the skin was included in the incision directly |
| 2 | 14 | indication for | indication | 4 | not applicable, slnb done |
| 3 | 1 | ncision and incision relation to tumor | incision and incision relation to tumor | 1 | overlying tumor |
| 4 | 3 | ncision in relation to tumor | incision in relation to tumor | 1 | not applicable mastectomy done |
| 5 | 1 | ndication | indication | 1 | primary treatment |
| 6 | 3 | ndication | indication | 1 | completion of mastectomy after initial bcs |

Appendix H: Autocorrected column names are automatically recorded and displayed
